# Clinicians’ Visual Attention During Suicide Screening Encounters: An Exploratory Eye-Tracking Study

**DOI:** 10.64898/2026.02.14.26346315

**Authors:** Doaa Alrefaei, Kaidi Huang, Ashwin Sukumar, Soussan Djamasbi, Bengisu Tulu, Rachel Davis Martin

## Abstract

Eye tracking is recognized as a gold standard for measuring visual attention and cognitive engagement. In this study, it offers a useful lens for understanding how primary care providers balance patient communication with navigation of electronic health records (EHRs). We used wearable eye tracking to collect visual information processing behavior and conducted a retrospective think-aloud protocol to examine how primary care clinicians processed suicide-related information (CAT-MH^®^) embedded in the EHR during simulated visits. Eye-movement data showed substantial visual attention directed toward the EHR, indicating added information-processing demands during communication. Retrospective think-aloud data supported the analysis of eye movement data by revealing that clinicians searched multiple record sections to verify risk indicators and often postponed suicide-related discussions until confirming relevant results. These findings illustrate how EHR-embedded screening tools shape clinical attention and encounter flow.

## 1 Introduction

Suicide is a major public health concern and remains a leading cause of preventable death worldwide (Lukaschek et al., 2024). In primary care, routine suicide risk screening is considered a core clinical responsibility because many individuals with suicidal ideation first present in this setting (Horowitz et al., 2023). As primary care providers frequently serve as the initial point of contact for patients with mental-health concerns, routine visits represent an important opportunity for early detection and intervention (Jeong et al., 2024).

High-quality assessment of suicide risk relies on standardized screening measures that can be integrated into electronic health record (EHR) systems. Computerized Adaptive Testing for Mental Health (CAT-MH®) is one such tool and has been used in primary care to assess suicide risk and related mental health concerns (Gibbons & deGruy, 2019). When integrated into the EHR, CAT-MH® results are available to clinicians during routine patient encounters (Chitavi et al., 2025).

Because suicide-related screening results are accessed during routine visits through the EHR, their usefulness depends on how clearly this information is presented and organized for clinicians (Ratwani et al., 2018). Prior work shows that clear EHR visualizations highlighting clinically meaningful information can reduce cognitive effort during clinical tasks and make it easier for providers to notice when patients may require urgent attention. (Pollack & Pratt, 2020).

Primary care encounters are cognitively demanding because providers manage multiple communication tasks at once. During routine visits, clinicians interact with patients while locating, reviewing, and interpreting information in the EHR, which requires continuous allocation of attention (Street et al., 2014).

Because vision is the dominant sense for sighted individuals, visual attention plays a central role in how information is accessed and processed during clinical work. Eye tracking provides an unobtrusive measure of visual attention and has become the gold standard for capturing how users allocate attention across tasks (Djamasbi, 2014). In this study, eye tracking is used to investigate how clinicians manage the complex interplay between patient communication and information engagement during routine primary care encounters.

Visual attention provides a useful lens for understanding how clinicians interact with patients and screens during encounters (Sqalli et al., 2023). However, far less is known about how primary care providers allocate visual attention when suicide risk indicators from digital screeners such as CAT-MH® are integrated into the EHR and available during routine visits. This represents an important interaction context to understand clinicians’ visual behavior during routine primary care encounters.

Because visual attention reflects cognitive effort, it can inform the design of systems that present information, such as suicide risk indicators, in ways that align with clinicians’ workflows. This, in turn, can reduce clinicians’ cognitive load during patient encounters, enabling them to attend more effectively to their patients. Therefore, in this study, we examine physicians’ visual attention during portions of the visit in which primary care providers are simultaneously interacting with the EHR and communicating with the patient, focusing on how embedded CAT-MH® outputs shape attention allocation and communication.

## 2 Background

### 2.1 Suicide Risk Assessments

Suicide risk assessment depends on clinicians’ access to and interpretation of displayed suicide-related information during routine encounters. Primary care providers frequently serve as the first point of contact for individuals experiencing suicidal ideation or related concerns (Jeong et al., 2024), which makes the way screening information is presented in the EHR clinically important. Computerized Adaptive Testing for Mental Health (CAT-MH®) supports this process by providing a digital adaptive testing system that measures multiple mental health constructs, including depression and suicide-related items (Gibbons et al., 2016; Gibbons & deGruy, 2019). When integrated into electronic health records (EHR), CAT-MH® provides structured scores or alerts that are available during the visit, allowing clinicians to review suicide-related information in real time and incorporate it into their communication with patients and clinical decision making (Gibbons & deGruy, 2019) without having to access an external report.

### 2.2 Clinician’s Cognitive Workload

Clinical encounters require providers to manage multiple concurrent tasks within a limited time frame. Routine visits involve frequent multitasking, task switching, and interruptions, which contribute to increased cognitive effort during the clinical workflow (Laxmisan et al., 2007). Interaction with patients often coincides with clinicians accessing and using the EHR, requiring clinicians to shift attention between listening, documenting, retrieving information, and clinical reasoning. These shifts place demand on cognitive resources and constrain providers’ capacity to engage simultaneously with multiple information sources.

EHR design influences these cognitive demands. Clinically relevant information is often distributed across multiple screens or sections, requiring navigation under time pressure (Ratwani et al., 2018; Walker et al., 2021). Prior work has linked EHR usability to excess cognitive workload and lower performance, while showing that more streamlined interfaces are associated with reduced workload and better task performance (Mazur et al., 2019). When suicide risk information is present in the EHR, screening results and alert information become part of clinicians’ ongoing workflows, where they may compete with other clinical demands and shape how and when risk-related information is attended to during patient care (Kline-Simon et al., 2020).

### 2.3 Eye Tracking for Measuring Clinical Attention

Because vision is the dominant sense for sighted individuals, visual attention plays a central role in how information is accessed and processed. Eye tracking provides an unobtrusive measure of visual attention and is widely used to study how users allocate attention across tasks and interfaces (Djamasbi, 2014). As such, eye tracking plays an increasingly important role in the design of human-centered solutions (Djamasbi, 2025).

In healthcare, eye tracking has been applied to examine how clinicians view electronic records, decision support tools, and other digital displays, and to evaluate the usability of EHR interfaces (King et al., 2017; Asan & Yang, 2015). Studies show that gaze patterns can reveal which parts of the record are fully read, which content is overlooked, and where users experience navigation difficulties (King et al., 2020; Brown et al., 2014).

Recent work also uses eye tracking to quantify aspects of cognitive load during interaction with clinical systems (Asgari et al., 2024; Kadhim et al., 2023). This line of research highlights that longer or more fixations on a specific screen of the EHR can reflect high cognitive demands during clinical workflow. In addition, recent work on face-to-face care, uses wearable eye-tracking devices to allow researchers capture how clinicians shift visual attention between patients and the EHR during real or simulated encounters (Jongerius et al., 2021; Jongerius et al., 2022). They show that clinicians repeatedly redirect gaze between the patient and screen as they move across tasks, such as history taking, documentation, and decision making (Jongerius et al., 2021; Jongerius et al., 2022). These shifts mark changes in conversational topics and moments when clinicians engage in more intensive information processing.

Research on clinical empathy and communication shows that a provider’s gaze toward the patient’s face should not be interpreted on its own as a sign of empathy or relationship quality, because clinicians often redirect their gaze to support cognitive tasks such as information retrieval and documentation, and patients interpret empathy within the broader interaction (Park et al., 2024; Nissan et al., 2026) These findings indicate that gaze patterns need to be understood in light of what the clinician is doing and discussing at that moment.

## 3 Methodology

### 3.1 Participants

Five primary care providers participated in this study (3 males, 2 females). All participants were practicing clinicians affiliated with a university-affiliated hospital in the northeastern United States, and each clinician completed one simulated routine primary care visit. Participants were recruited through internal clinical networks. Trained individuals served as standardized patients to simulate realistic clinical encounters in this study. All procedures were approved by the institutional review board (IRB).

### 3.2 Apparatus

#### Eye-tracking glasses

Providers’ eye movements toward both the patient and the EHR were recorded using Argus Science wearable eye-tracking glasses, sampling at 120 Hz. A velocity threshold of 30°/s was used to determine saccade onset, and minimum fixation duration was set to 100 ms (Holmqvist et al., 2011). The eye-tracking system includes one inward-facing camera per eye and an outward-facing scene camera.

#### Device configuration

The eye-tracking device was connected to a laptop that served as the recording station. A second laptop was placed on a movable desk and used by participating providers to access and navigate the EHR during the encounter (see Fig 1). This setup enabled continuous and synchronized recording of providers’ gaze behavior both while interacting with the EHR and while attending to the patient during clinician–patient communication throughout the visit. The Argus ET-Vision Version 1.0.7.6 captured eye movement data. After each recording, the software produced a file called ‘scene video,’ which overlaid the gaze stream (collected with the internalfacing camera of the wearable eye tracker) onto the video captured by the external-facing camera of the wearable eye tracker (visual field of the participants). The Argus ET-Analysis Version 1.1.2.6 was used to extract eye movement metrics reported for this analysis from the scene video.

**Fig. 1.**
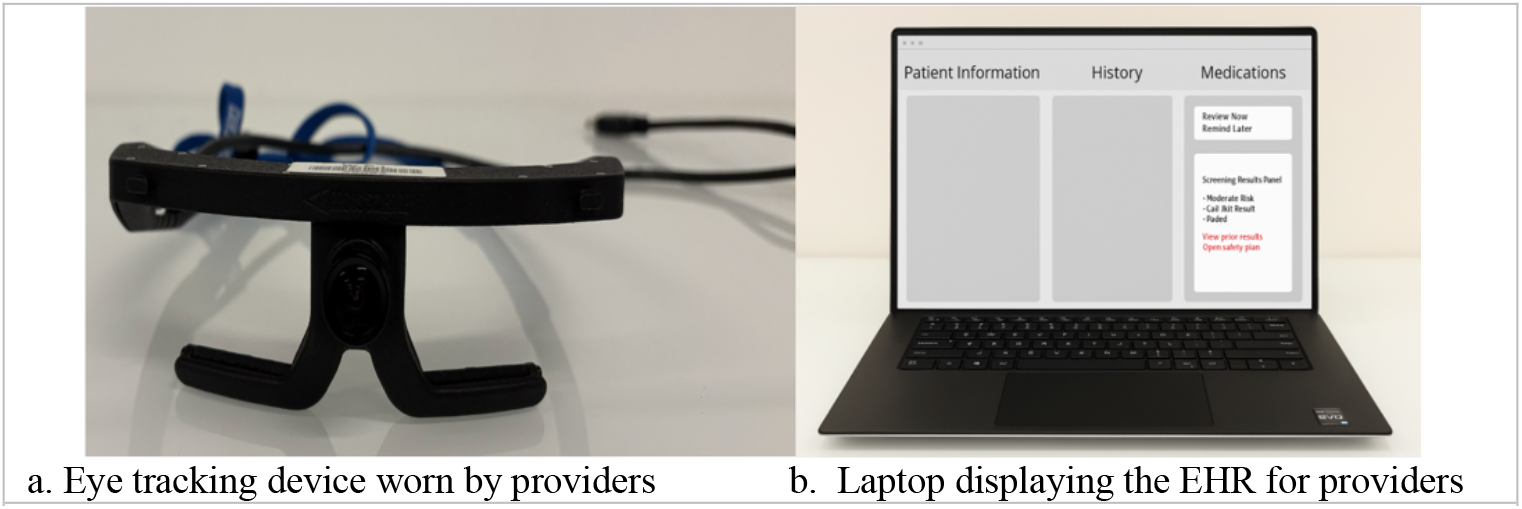
Hardware set up for simulated clinical encounter.

#### Calibration

Before data collection, a calibration procedure was performed individually for each provider using a single-point display to align gaze estimates to the scene camera. Calibration and scene camera settings (rotation, focus) were adjusted as needed per participant to ensure accurate tracking.

### 3.3 Task and Data Collection

#### Simulation environment

The study took place in a simulated primary care examination room designed to resemble a standard outpatient setting. Before starting the data collection, providers were invited to adjust the position of the movable desk and the laptop to accommodate their typical workflow, after which eye-tracker calibration was performed. The encounter followed a typical primary care sequence, including the clinician entering the room, engaging in conversation with the patient, reviewing information in the EHR, and documenting care (see Fig. 2).

**Fig. 2.**
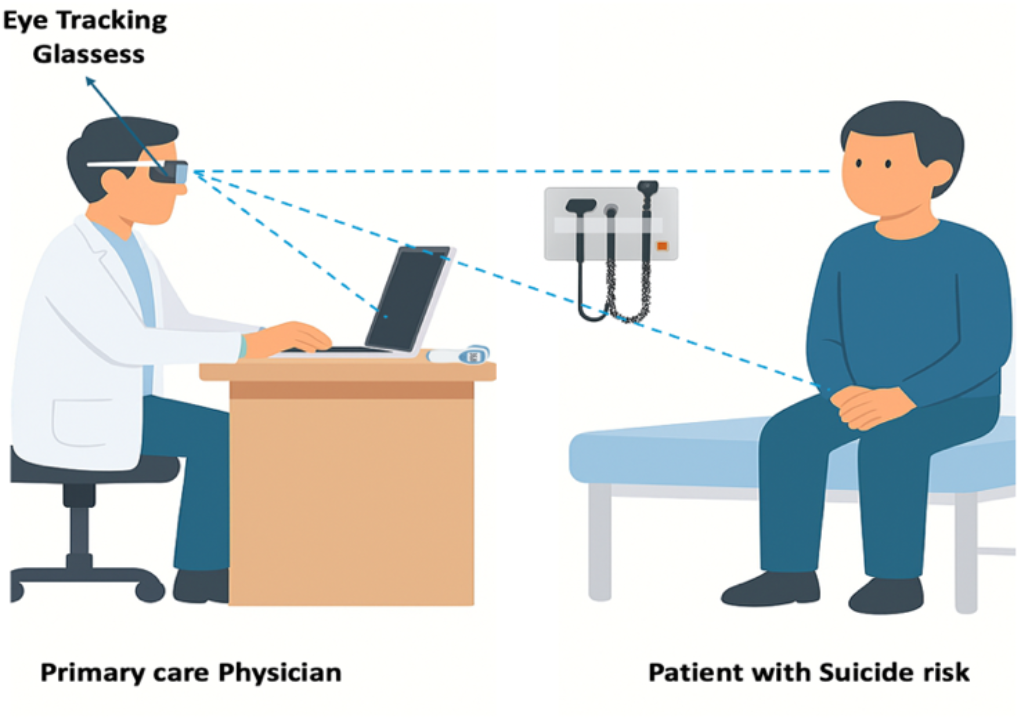
physician wearing wearable eye-tracking data during Simulated routine primary care visit

#### Simulated encounter

Providers were instructed to conduct a routine primary care visit with a standardized patient. Each clinician was assigned a scenario case representing a new patient encounter, during which the patient communicated symptoms and concerns verbally. The patient’s EHR included CAT-MH^®^ suicide risk results, which were available during the encounter. CAT-MH^®^ outputs were presented as interruptive pop-up alerts and were also accessible within the record for later review (See Fig. 3)

**Fig. 3.**
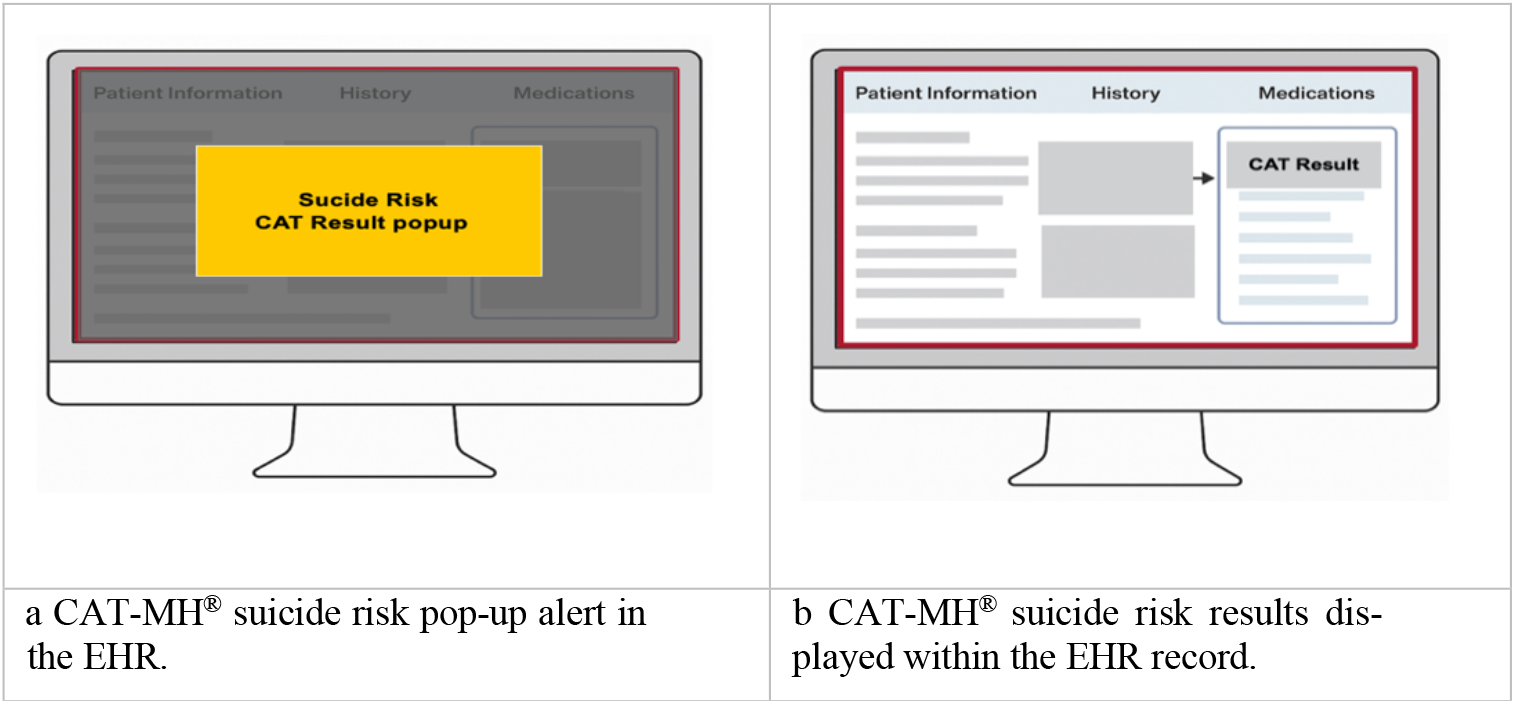
physician wearing wearable eye-tracking data during Simulated routine primary care visit

#### Retrospective think-aloud

Immediately after completing the simulated visit, Providers participated in a gaze-cued retrospective think-aloud session (Elling et al., 2012; Alrefaei et al., 2023). During this session, clinicians viewed synchronized gaze replays of their encounter and were asked to recall their thought processes while reviewing suicide-related outputs in the EHR. This procedure elicited reflective comments on navigation strategies, interpretation of CAT-MH^®^ results, and timing of suicide-related communication.

#### Analysis

The analysis of the eye movements reported in this study focuses on segments of the encounter in which providers were simultaneously interacting with the EHR and communicating with the patient. Phases other than EHR interaction and patient communication (e.g., portions of the visit involving physical examination) were excluded from the analysis.

### 3.4 Measure

#### Areas of Interest (AOIs)

To examine eye movement behavior, the first step is to specify the regions of the visual field (commonly known as areas of interest or AOIs) on which eye movement analysis will be conducted (Djamasbi, 2014). For this study, two AOIs were delineated on the scene video: (1) a Laptop AOI, encompassing the computer used by clinicians to review patient records, and (2) a Patient AOI, encompassing the patient’s entire body. These AOIs enabled us to examine the distribution of providers’ visual attention during communication-intensive segments of the clinical encounter. Gaze outside these AOIs (e.g., clinical environment or physical examination) was excluded from the analysis reported in this paper.

#### Eye-movement Metrics

Three eye-movement metrics were used to quantify visual attention in this study: visit duration, fixation count, and fixation duration. Visit duration refers to the amount of time spent viewing an AOI. Fixation count refers to the number of discrete fixations. Fixation duration refers to the duration of a fixation. These metrics are commonly used in eye-tracking research to assess visual attention and cognitive effort (Holmqvist et al., 2011; Poole & Ball, 2006).

Because encounter durations varied across providers, all eye-tracking metrics were time-normalized (Alrefaei et al., 2023). Relative visit duration for each AOI was computed by dividing the AOI-specific visit duration by the cumulative visit duration across both AOIs. Similarly, relative fixation count and relative fixation duration were calculated by dividing each metric by its corresponding cumulative value across both AOIs.

## 4 Result

### 4.1 Findings from Eye movement metrics

Figure 4 displays eye-tracking metrics that quantify providers’ cognitive effort while communicating with patients and interacting with the EHR. Clinicians allocated a greater proportion of their time reviewing the patient information that was provided through the laptop (53%) compared to placing their gaze toward the patient (47%), indicating higher visual and cognitive engagement with the EHR during the encounters. Figure 4 also shows that relative fixation count (FC) was higher for the laptop (56.63%) than for the patient (43.36%). A similar pattern was observed for relative fixation duration (FD), with 57.78% of fixation time directed toward the laptop and 42.21% toward the patient. The distribution of viewing behavior revealed by the above analysis suggests that EHR interaction demanded a substantial share of providers’ attentional resources during patient communication.

**Fig. 4.**
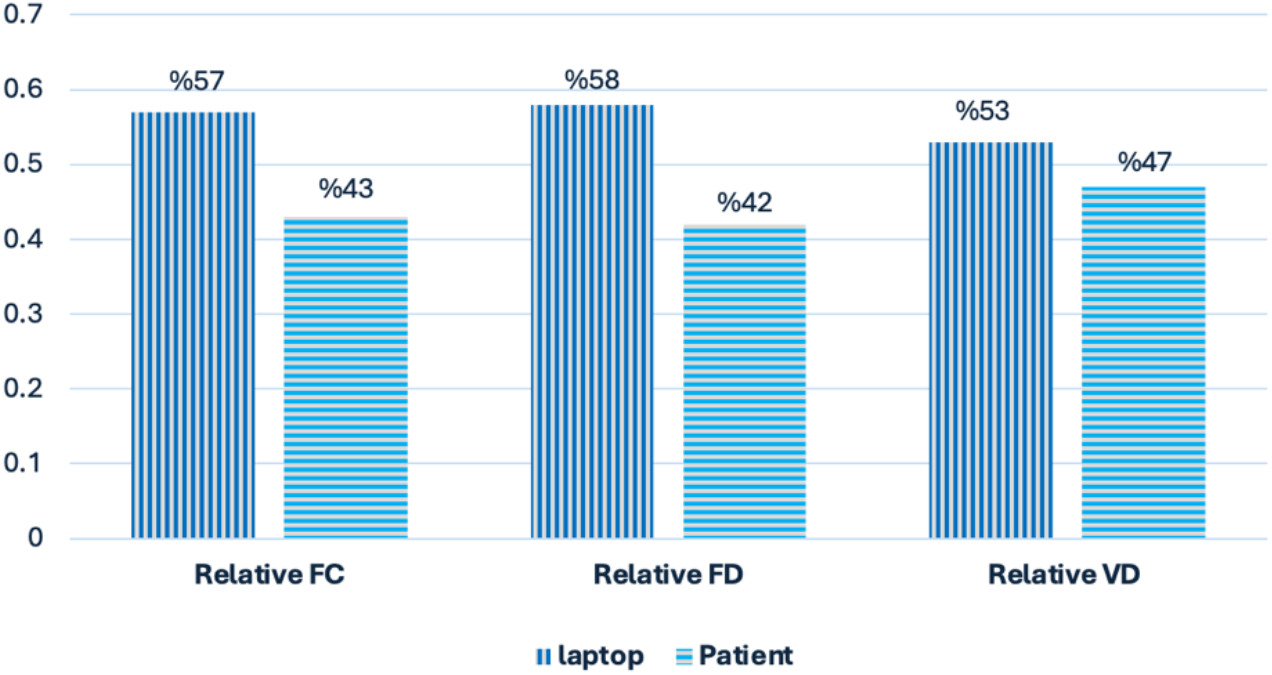
Eye movements for laptop and patient

### 4.2 Findings from the Retrospective Think Aloud Protocol

The retrospective think-aloud sessions indicated that physicians devoted considerable portions of the encounter to viewing and navigating the EHR because they were actively locating and interpreting CAT-MH^®^ outputs. Participating physicians reported scanning multiple sections of the chart for screening results, verifying whether suicide-related indicators were present, and interpreting alert content in the context of the visit. This often required switching between different EHR views, including flow-sheets and alert panels, before re-engaging with the patient.

Providers noted that locating CAT-MH^®^ results was not always straightforward, especially when they were unsure where information was stored or whether additional indicators existed elsewhere in the chart. When results did not appear in the anticipated locations, clinicians continued searching or temporarily paused the interaction with the patient to confirm whether a risk signal was present. These behaviors were evident in the scene video, which overlays providers’ gaze behavior onto their field of view, revealing how providers shifted attention to the laptop screen to interpret or act on the information.

Navigation demands influenced communication timing during the encounter. Providers delayed mental-health discussions until they had verified relevant indicators in the EHR, particularly suicide-related results that could require immediate follow-up, documentation, or adjustments to the visit agenda, despite the results and the notification pop-up during their navigation. Clinicians also emphasized that screening outputs are most useful when available before entering the exam room, because this allows them to plan the visit and decide when and how to address mental-health concerns without receiving unexpected alerts mid-visit.

The think-aloud protocol, therefore, highlighted how CAT-MH^®^ outputs influenced encounter flow by increasing time spent navigating the EHR, shaping the timing of sensitive conversations, and prompting verification of automated assessments before clinical communication.

## 5 Discussion

In this study, we examined how primary care providers allocated visual attention and processed suicide-related screening information during simulated primary care encounters in which CAT-MH^®^ outputs were available within the EHR. We combined the analysis of eye movement data with a retrospective think-aloud protocol to capture both observable gaze behavior and clinicians’ reflections on how suicide-related information was located, interpreted, and used while communication with the patient was ongoing.

Eye-movement data showed that clinicians devoted more visual attention to the EHR during segments of the encounter that involved both communication and information retrieval. This is not surprising, as clinicians need to review patient information to conduct a productive visit and make effective treatment decisions. For example, feed-back collected during the retrospective think-aloud sessions indicated that providers searched the chart for CAT-MH^®^ outputs to confirm the presence or absence of suicide-related indicators and to determine how to interpret these outputs within the context of the encounter. This pattern suggests that the presentation of CAT-MH^®^ outputs within the EHR introduced additional information-processing demands. Locating, verifying, and interpreting suicide-related information naturally contributed to increased cognitive effort during the encounter.

Clinicians also reported that locating CAT-MH^®^ outputs was not always straightforward and that they had to scan multiple sections of the EHR to verify suicide screening results. The increase in cognitive demand observed in our study is consistent with the findings of prior research that show distributed EHR information increases clinicians’ cognitive load during primary care visits (Ratwani et al., 2018; Walker et al., 2021)

In addition to increasing providers’ cognitive load, the distribution of suicide-related information within the EHR affected the flow of communication with patients, as providers reported delaying discussions of suicide-related topics until they had confirmed relevant indicators in the medical record. Taken together, these findings suggest that redesigning the placement and labeling of suicide-related information (e.g., through clear placement, clear labeling, and minimal navigation) can not only reduce clinicians’ cognitive effort but also improve physician–patient communication during the visit.

## 6 Strengths and Limitation

As with any study, our investigation has both strengths and limitations. A key strength of this work is the use of eye-tracking goggles to examine clinicians’ access to suicide-related information in the EHR during a simulated outpatient encounter. Although screen-based eye trackers could capture clinicians’ viewing behavior on the EHR, they would not allow observation of how physicians balance attention between the patient and the screen while processing EHR information. Capturing attention distribution in multitasking environments enables a more nuanced understanding of clinician behavior and supports the design of systems that help providers process relevant information more effectively. It also allows examination of attention allocation in relation to the clinician’s ongoing activities during the encounter.

A limitation of this study is the small number of cases analyzed. However, data collection is ongoing, and the expanded dataset will allow us to validate these findings and develop design recommendations based on a larger sample.

## 7 Conclusion

This study advances human-centered design research by demonstrating the practical value of integrating eye-tracking data into the product development process. By linking objective measures of visual attention with clinicians’ self-reported experiences, we were able to generate meaningful design insights, even with a modest sample size, that support efficient information processing, informed clinical decision making, and effective patient-centered communication.

In addition, this work makes a methodological contribution to human-centered design by employing mobile eye tracking to examine attention distribution in multitasking environments. Unlike traditional screen-based eye trackers, which capture only on-screen gaze behavior, mobile eye tracking enables the assessment of visual attention across the entire visual environment. This approach provides a more comprehensive understanding of how individuals allocate attention among competing objects and tasks. By overcoming a key limitation of conventional screen-based eye-tracking techniques, our study offers a more ecologically valid perspective on information-use behaviors in information-rich settings and opens new opportunities for designing technologies that better support the cognitive and communicative demands of multitasking situations, such as clinicians’ use of EHRs during outpatient visits.

## Data Availability

All data produced in the present work are contained in the manuscript

## Acknowledgments

Research reported in this publication was supported by the National Institute Of Mental Health of the National Institutes of Health under Award Number P50MH129701. The content is solely the responsibility of the authors and does not necessarily represent the official views of the National Institutes of Health

